# Predicting Mental and Psychomotor Delay in Very Pre-term Infants using Large Language Models

**DOI:** 10.1101/2025.07.31.25332524

**Authors:** Zhewen Huang, Michael J. Flory, Phyllis M. Kittler, Ha T. T. Phan, Gözde M. Demirci, Anne D. Gordon, Santosh M. Parab, Chia-Ling Tsai

## Abstract

Very preterm infants face a considerably higher risk of neurodevelopmental delays, making early diagnosis and timely intervention crucial for improving long-term outcomes. In this study, we utilized large language models (LLMs) to predict mental and psychomotor delays at 25 months using maternal and perinatal records combined with longitudinal features up to 22 months of age. The LLMs were employed to generate natural language descriptions for each infant, which were then used as input for a language model-based classifier to perform predictions. Our model achieved a 4.2% increase in AUCROC in mental delay prediction and 3.2% increase in psychomotor delay prediction 3 months before the 25-month assessment, compared to a random forest-based model for numerical tabular data only. These findings highlight the potential of LLMs as powerful tools for assessing the risk of neurodevelopmental delays in preterm infants.

## Introduction

Preterm birth is highly prevalent, affecting nearly 1 in 10 infants [1]. The risk of mental and psychomotor delays in preterm infants remains high despite the substantial advances in medical care [2]. The risk is even higher for very preterm infants, born earlier than 34 weeks. In 2022, 2.78% of the infants were very preterm infants in the United States [3]. Families and society dedicate considerable emotional and financial resources to support these infants. The annual societal economic cost associated with preterm birth in the United States is estimated $25.2B [4]. Early identification and timely intervention of developmental delays in very preterm infants are essential to optimize outcomes and unlock the full potential of affected children [5].

Various machine learning approaches have focused on predicting the outcome of preterm infants. A comparative study of multiple machine learning models, including logistic regression, random forest, support vector machines, and gradient boosting machines, evaluated their performance in predicting 2-year cognitive outcomes in very preterm infants [6]. A random forest model [7] was employed to predict mortality in preterm infants, demonstrating superior performance compared to the widely used Clinical Risk Index for Babies (CRIB). DeepPBSMonitor [8], a deep learning model integrates real-time vital sign data and static clinical variables to predict mortality risk during NICU hospitalization. These studies are limited to numerical tabular data, which restricts their broader application to medical prediction tasks involving valuable categorical and textual information [9].

Language models designed to predict the likelihood of future or missing tokens in a sequence are particularly well-suited for analyzing medical data, which often includes substantial textual information like medical history. Additionally, natural language enables the incorporation of domain-specific information based on feature names to supplement description on values in tabular data, which is overlooked by traditional machine learning methods. Pretrained language models enable high downstream task performance even with relatively small datasets, as they carry rich domain-specific knowledge learned from large-scale pretraining. This helps boost performance beyond what the task-specific data alone can offer. As a result, there are already many applications of language models in the medical field, for example, Med-BERT [10], a BERT based model pretrained on diagnostic information from 20 million patients. MeDeBERTa [11], another BERT based model, is pretrained on electronic health records.

Recently, large language models (LLMs) have experienced rapid development, including larger parameter counts compared to traditional language models such as BERT. The advanced capabilities of LLMs, including in-context learning, instruction following, and multi-step reasoning, enable them to capture intricate inter-feature logic and domain-specific knowledge, making them highly suitable for predictive tasks in the medical field. LLMs achieve better results for multiple tasks in the medical field than language models. GatorTron [12], an LLM with 8.9 billion parameters, outperforms transformer-based language models in predicting outcomes from electronic health records. Med-PaLM 2 [13] achieved a 50% higher score on the MedQA dataset [14], which includes multiple choice medical questions in the clinical field, compared to language models with fewer parameters like PubMedBERT [15]. Med-PaLM 2 also surpassed the performance of general-purpose LLMs, such as GPT-3.5 [16].

Despite these advancements, limited studies utilizing LLMs focus on addressing the problems of preterm infant neurodevelopment, such as identifying the risk of neurodevelopmental delays of very preterm infants. The goal of this study is to apply the LLMs to predict the mental and psychomotor delays at 25 months of age in very preterm infants, combining the tabular data of clinically available variables from the perinatal period and longitudinal neurodevelopmental assessments.

## Methods

Our dataset reflects the inherent class imbalance of the conditions examined, with most preterm infants classified as having no developmental delay. Additionally, like many longitudinal studies, our dataset contains a considerable percentage of incomplete records, even after removing sparse variables. This is primarily due to families missing follow-up visits or losing contact with the research team.

Consequently, robust data preprocessing of the tabular data is essential. Our key steps are targeted data selection, separate imputation of missing values for training and testing sets, and augmentation of the minority class within the training data. Each infant’s tabular data is then transformed into a natural language description using LLMs, which serves as input for our prediction model.

### Data preprocessing

The original de-identified dataset came from a study cohort containing 3,567 infants recruited from Richmond University Medical Center level III NICU based on previously reported criteria [17]. We selected all infants with a gestational age of ≤33 weeks, resulting in a reduced dataset of 1,109 infants for this study. There are records of multiple visits for each infant, providing longitudinal data samples over time. To enhance the representativeness of the dataset, we excluded variables with more than 80% missing values and those measured beyond 25 months. The steps for data selection are illustrated in Figure 1.

**Figure 1.**
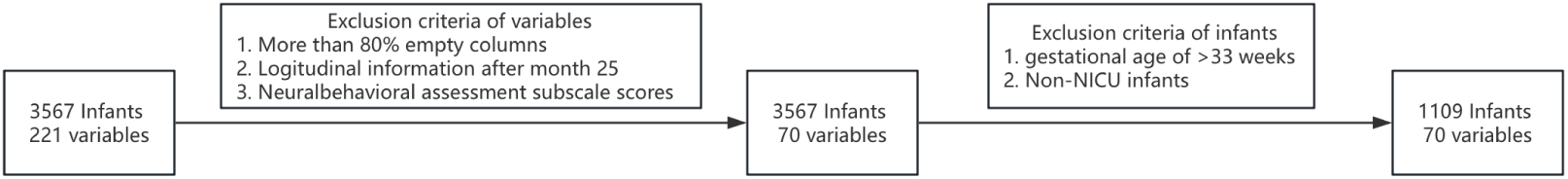
Data selection.

Imputing missing data is critical for this model because we need to maximize data utilization. A random forest-based imputation method was employed to handle missing values, as it is well-suited for datasets with diverse data types. The missing imputation model was trained only on the training set before applying it to both training and testing sets, ensuring that no information from the test set was utilized during the imputation process. Importantly, no target values were missing in the original test set, preserving its authenticity for model evaluation.

Moreover, there was a class imbalance among participants with only 15% of infants classified as delayed at 25 months of age after imputing missing target values. Imbalanced data can cause the prediction model to overpredict outcomes for the majority class [18]. To address this issue and improve the robustness of the prediction model, the Synthetic Minority Oversampling Technique (SMOTE) [19] was applied to generate additional samples for the minority class. Importantly, SMOTE was performed exclusively on the training set to ensure that the test set remained unaltered and representative of the original data. Figure 2 illustrates the overall steps of data preprocessing.

**Figure 2.**
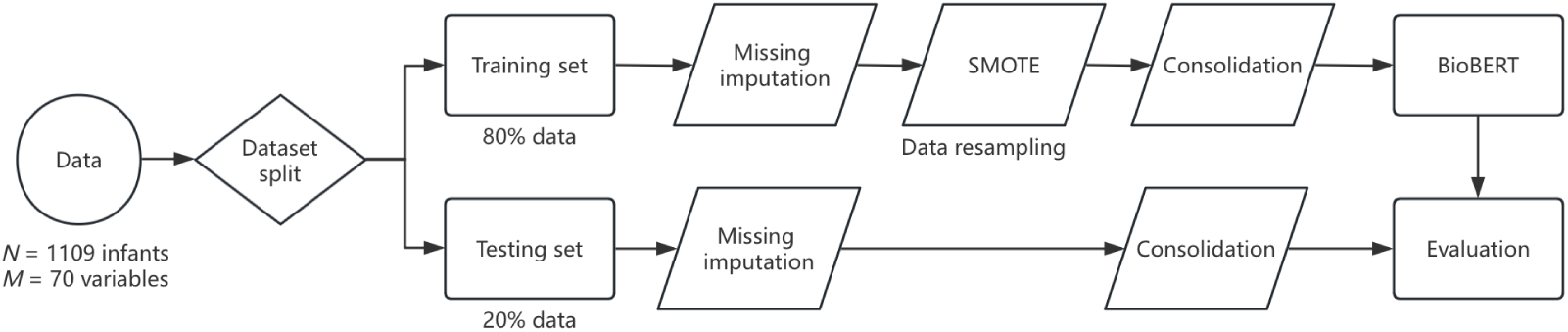
Data preprocessing flowchart.

### Data consolidation

In the original dataset, many feature names and data values were vague and challenging to interpret, both for laypersons and the LLM, as shown in the example.

| abnhus | placenta | steroids | apnea | asphyxiahypoxia | austim | austim_sib | phenobrb | csection |
| --- | --- | --- | --- | --- | --- | --- | --- | --- |
| 0 | 1 | 1 | 0 | 0 | 0 | 0 | 0 | 1 |

To address this, we transformed each row into natural language. First, we excluded zero values from binary columns–such features were not observed–to help minimize the influence of hallucinations during the LLM’s generation process and focus on the important features for each infant [20]. Second, we changed the feature names and numerical values into natural language. Specifically, we concatenated the feature names with their corresponding cell values to create key-value pairs, separated by semicolons. In the preceding example, the binary columns **apnea, asphyxiahypoxia, austim, austim_sib**, and **phenobrb** contain only zero values and are thus excluded from the paraphrase, **abnhus** is a multi-class label, not a binary attribute, and is kept. For the remaining features, a value of zero in the **abnhus** column indicates a head ultrasound shows the normal result and a value of one in the **placenta** column means an abnormal placenta. Consequently, the linearization of this example is as follows:

~~~
head ultrasound result normal; abnormal placenta; administered steroids;
baby delivered via c-section
~~~

We leveraged LLMs to enrich the linearization, utilizing GPT-3.5 for generating paraphrases. To guide GPT-3.5, we used a one-shot approach, providing an example output generated by GPT-4o [21] from a training set linearization. Furthermore, we included specific instructions emphasizing medical causal order and a step-by-step thought process. These additions were crucial for enabling the LLM to better understand the intricate relationships among different medical features and retain essential information vital for accurate medical prediction. A simple prompt of providing only the linearization and instructing the LLM to paraphrase often fails to capture the underlying logic, resulting in lower consistency in output quality and style [22]. The prompt we used is as follows:

~~~
{Example}
Original: {linearization}
Paraphrased:
Please paraphrase in Medical Causal Order like the example before. Give ONE paragraph without annotations. Include all digits in Bayley Scales scores (mental & motor) and Neurobehavioral total abnormality. Think step-by-step before outputting.
~~~

Comparing to the simpler prompt, the paraphrase generated with our detailed prompt shows improved logical flow between features, better temporal ordering, more descriptive trends and necessary details, and enhanced readability. This evidence strongly suggests that our prompting approach, incorporating targeted instructions and an illustrative example, is highly effective in guiding the LLM to comprehend the underlying medical logic and generate better paraphrases.

Additionally, data augmentation using LLMs offers substantial potential, particularly for relatively small datasets like ours. To further enhance the dataset, we generated five different paraphrased entries for each row in the dataset, which is helpful to address the pervasive issue of limited data volume. The generated paraphrases transform the key-value pairs into richer natural language descriptions, effectively capturing the inter-feature logic and incorporating valuable domain-specific knowledge. These enhanced descriptions were then paired with the target variables to serve as input for our prediction model.

### Prediction model

The prediction model employed in this study is BioBERT [23], a BERT based model pretrained on extensive biomedical corpora containing 20 billion words. Compared to the standard BERT model, BioBERT is specifically fine-tuned for the biomedical domain, allowing it to capture domain-specific knowledge and achieve superior performance in biomedical text mining tasks. Given the abundance of biomedical text in our dataset, BioBERT is particularly well-suited for this study.

The paraphrased description of an infant serves as the input of the BioBERT based prediction model. The model outputs a prediction indicating whether the infant has mental/psychomotor delay at 25 months: 0 indicates that the infant has no delay, 1 indicates that the infant has delay.

## Results

### Dataset

We utilized a published de-identified dataset of 1,109 very preterm infants recruited from Richmond University Medical Center level III NICU [17]. Table 1 shows some most important demographic and clinical characteristics of the study population in our dataset according to the analysis in [17]. This dataset includes maternal, perinatal, and longitudinal records:

**Table 1:**
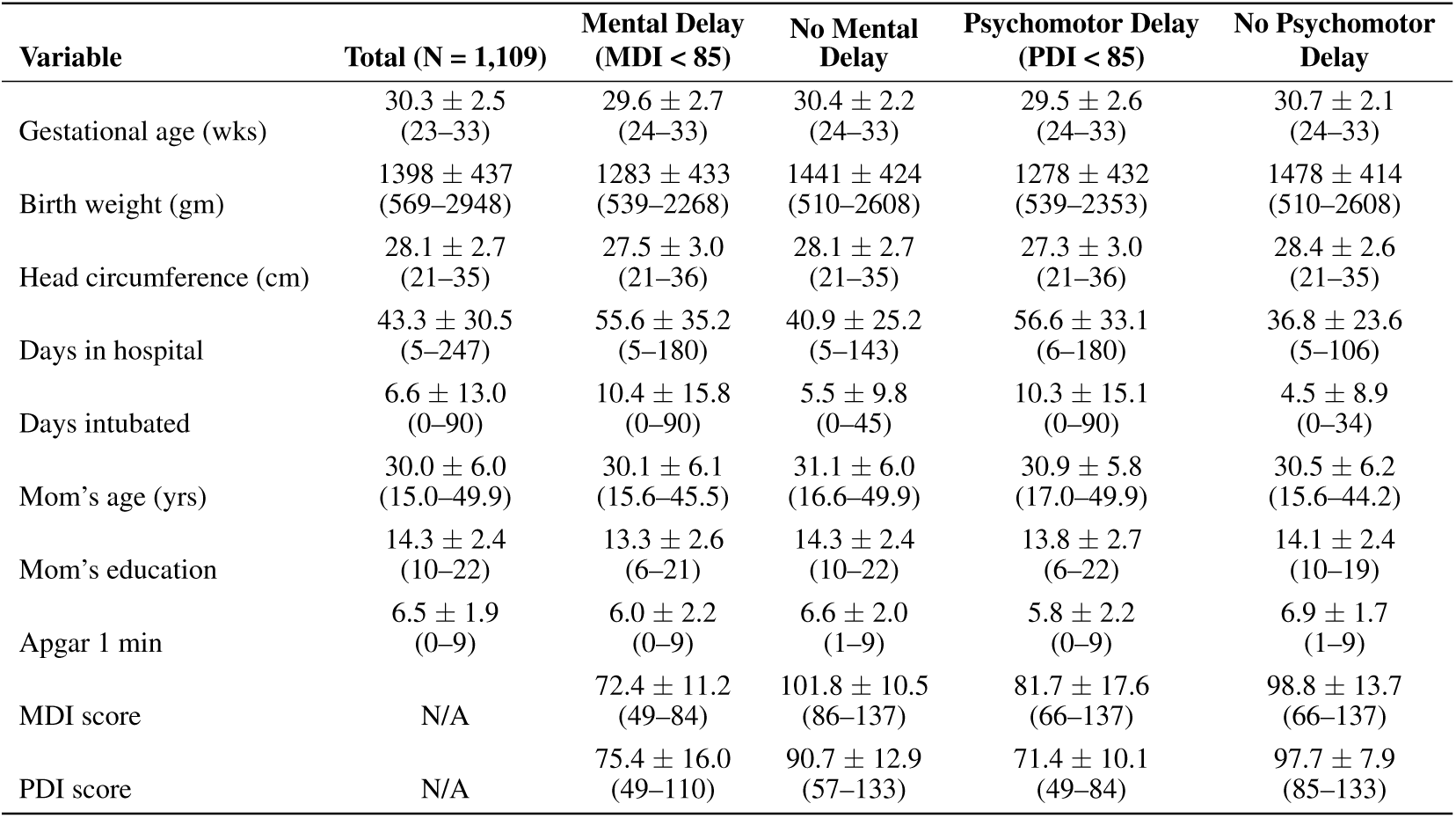
Demographic and clinical characteristics of the study population.

- At birth: The features measured at birth include a comprehensive set of variables related to the mother’s health and perinatal information. Additionally, they extensively cover maternal demographic information and substance exposure. They also cover the infant’s postnatal status, body measurements, and neonatal health issues and treatments.
- Rapid Neonatal Neurobehavioral Assessment (RNNA): RNNA [24] assesses neurobehavioral functions at two time points: at birth and one month of age. Designed specifically for high-risk neonates, the RNNA score has been validated as a reliable predictor of future mental and motor performance. Higher scores indicating a greater degree of abnormality. For better understanding by language models, we denoted it as neurobehavioral total abnormality.
- Bayley Scales of Infant and Toddler Development (BSID): BSIDs measure infants’ mental and psychomotor development and are divided into two components. The Mental Developmental Index (MDI) score evaluates cognitive and language abilities, the Psychomotor Developmental Index (PDI) score assesses fine and gross motor skills [25, 26]. BSIDs were conducted at 4, 7, 10, 13, 16, 19, 22, and 25 months.

Since our goal was to predict neurodevelopmental delays as far into the future as possible, we used the 25-month BSID assessment as the target outcome. The standard population mean for both MDI and PDI scores is 100, with a standard deviation of 15. Therefore, for this study, scores below 85 were classified as indicative of developmental delay.

### Prediction models and performance measurement

The predictor variables, as summarized in Table 2, are used in two separate analyses: one for predicting delays in MDI and another in PDI [17]. Several models were devised to study various combinations of the predictor variables. Model 1 includes only variables measured at birth. Model 2 adds to Model 1 the RNNA measured at birth and one month, indicating the development of infants in the first month. In the consecutive models, Model 3 to 9, the longitudinal follow-up Bayley scores for the corresponding month are added to Model 2, resulting in 9 separate models with two target values each.

**Table 2:**
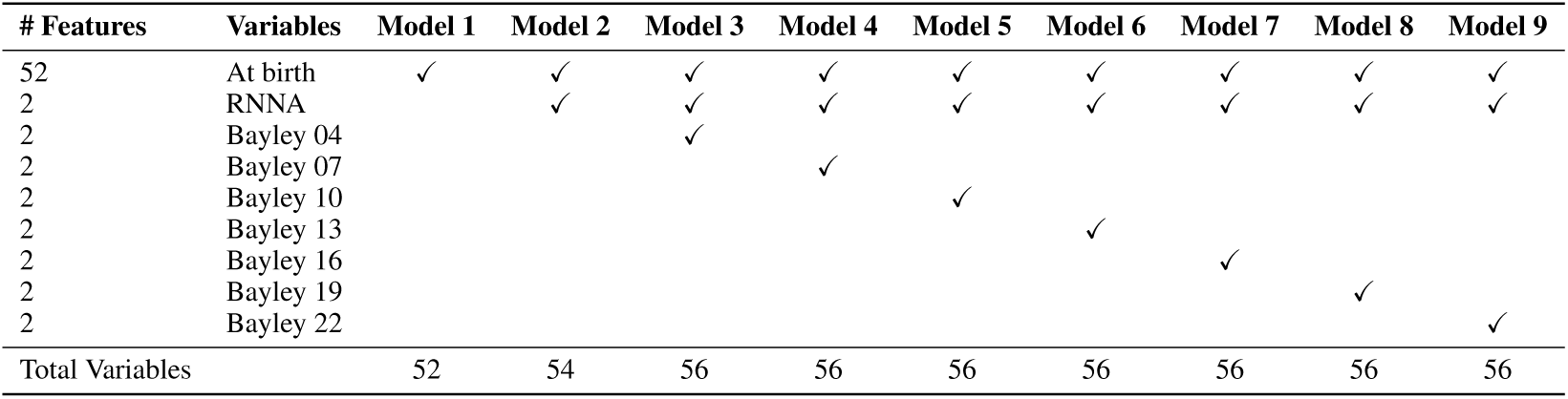
Predictor information for Model 1–9.

The first set of performance metrics includes sensitivity, specificity, and balanced accuracy. These metrics are particularly useful for evaluating model performance on imbalanced datasets. In our study, true positives represent delayed infants that are correctly predicted by the model. Sensitivity quantifies the model’s ability to predict delayed infants, specificity measures its ability to predict non-delayed infants. The balanced accuracy is the average of sensitivity and specificity.

AUCROC, Area Under the Receiver Operating Characteristic (ROC) curve, summarizes the trade-off between the true positive rate and the false positive rate. It is a widely used metric for evaluating the performance of binary classification models, especially for imbalanced datasets. By changing the threshold, the ROC curve offers an integrative view of model performance.

Considering only 52 maternal and perinatal variables measured at birth, Table 3 shows the balanced accuracy between Model 1 and common machine learning models for tabular data. Our model achieved the best performance when predicting both the MDI and PDI targets. Among the traditional machine learning models, the random forest model outperformed the others. It demonstrated a substantial advantage in predicting the MDI target, which outweighed its slightly lower performance compared to AdaBoost and XGBoost on the PDI target. Therefore, we selected the random forest model as the representative traditional machine learning model for subsequent experiments.

**Table 3:** Comparison of performance on predicting mental and psychomotor development using maternal and perinatal records (with Model 1 and machine learning models).

| Model | Mental Development (MDI) |  | Psychomotor Development (PDI) |  |
| --- | --- | --- | --- | --- |
|  | Balanced Accuracy | AUCROC | Balanced Accuracy | AUCROC |
| **Logistic Regression** | 60.3% | 0.676 | 61.9% | 0.670 |
| **GaussianNB** | 63.0% | 0.675 | 60.1% | 0.661 |
| **SVM (Linear)** | 60.7% | 0.687 | 63.2% | 0.669 |
| **SVM (Poly)** | 59.6% | 0.618 | 61.1% | 0.666 |
| **AdaBoost** | 62.9% | 0.675 | 63.5% | 0.724 |
| **XGBoost** | 62.4% | 0.712 | 62.6% | 0.720 |
| **Random Forest** | 63.9% | 0.732 | 62.2% | 0.722 |
| **LLMs** | **65.1%** | **0.748** | **63.7%** | **0.726** |

Highest values are emphasized in bold

The result of incorporating longitudinal data into the modeling process is summarized in Table 4 for both MDI and PDI target variables. Our model demonstrates superior performance over the random forest in most of the models. Furthermore, we observe that Model 9, which includes MDI and PDI scores measured at 22 months of age, performs the best.

**Table 4:**
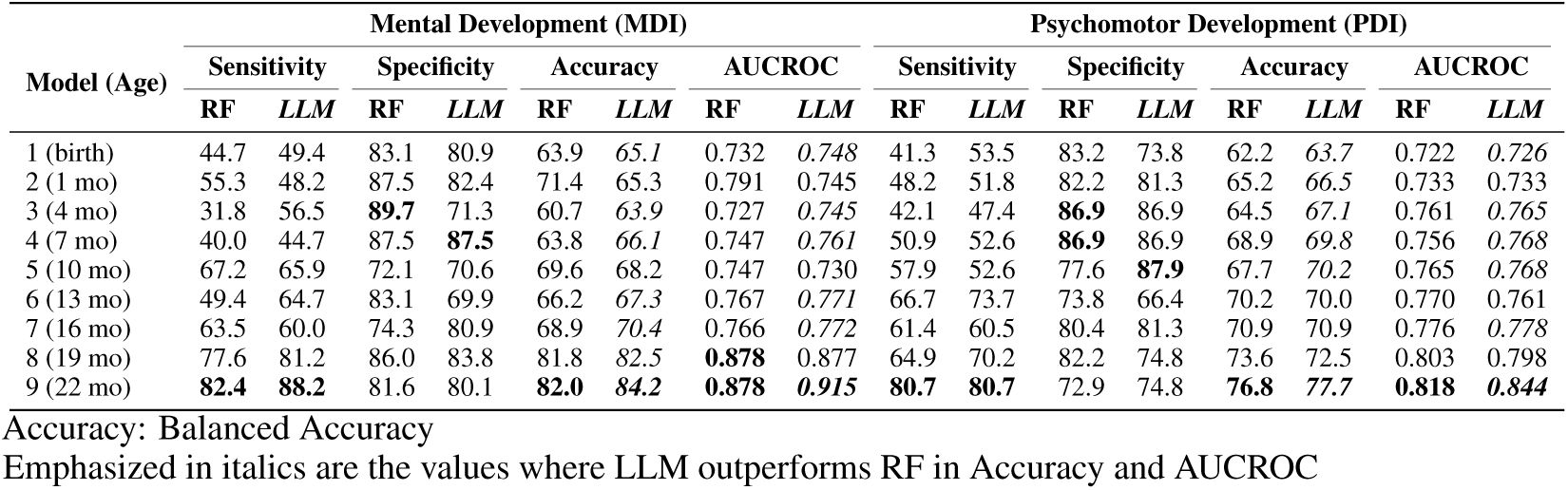
Model accuracies for target value of mental and psychomotor delay at month 25.

| Model (Age) | Mental Development (MDI) |  |  |  |  |  |  |  | Psychomotor Development (PDI) |  |  |  |  |  |  |  |
| --- | --- | --- | --- | --- | --- | --- | --- | --- | --- | --- | --- | --- | --- | --- | --- | --- |
|  | Sensitivity |  | Specificity |  | Accuracy |  | AUCROC |  | Sensitivity |  | Specificity |  | Accuracy |  | AUCROC |  |
|  | RF | LLM | RF | LLM | RF | LLM | RF | LLM | RF | LLM | RF | LLM | RF | LLM | RF | LLM |
| 1 (birth) | 44.7 | 49.4 | 83.1 | 80.9 | 63.9 | *65.1* | 0.732 | *0.748* | 41.3 | 53.5 | 83.2 | 73.8 | 62.2 | *63.7* | 0.722 | *0.726* |
| 2 (1 mo) | 55.3 | 48.2 | 87.5 | 82.4 | 71.4 | 65.3 | 0.791 | 0.745 | 48.2 | 51.8 | 82.2 | 81.3 | 65.2 | *66.5* | 0.733 | 0.733 |
| 3 (4 mo) | 31.8 | 56.5 | **89.7** | 71.3 | 60.7 | *63.9* | 0.727 | *0.745* | 42.1 | 47.4 | **86.9** | 86.9 | 64.5 | *67.1* | 0.761 | *0.765* |
| 4 (7 mo) | 40.0 | 44.7 | 87.5 | **87.5** | 63.8 | *66.1* | 0.747 | *0.761* | 50.9 | 52.6 | **86.9** | 86.9 | 68.9 | *69.8* | 0.756 | *0.768* |
| 5 (10 mo) | 67.2 | 65.9 | 72.1 | 70.6 | 69.6 | 68.2 | 0.747 | 0.730 | 57.9 | 52.6 | 77.6 | **87.9** | 67.7 | *70.2* | 0.765 | *0.768* |
| 6 (13 mo) | 49.4 | 64.7 | 83.1 | 69.9 | 66.2 | *67.3* | 0.767 | *0.771* | 66.7 | 73.7 | 73.8 | 66.4 | 70.2 | 70.0 | 0.770 | 0.761 |
| 7 (16 mo) | 63.5 | 60.0 | 74.3 | 80.9 | 68.9 | *70.4* | 0.766 | *0.772* | 61.4 | 60.5 | 80.4 | 81.3 | 70.9 | 70.9 | 0.776 | *0.778* |
| 8 (19 mo) | 77.6 | 81.2 | 86.0 | 83.8 | 81.8 | *82.5* | **0.878** | 0.877 | 64.9 | 70.2 | 82.2 | 74.8 | 73.6 | 72.5 | 0.803 | 0.798 |
| 9 (22 mo) | **82.4** | **88.2** | 81.6 | 80.1 | **82.0** | *84.2* | **0.878** | *0.915* | **80.7** | **80.7** | 72.9 | 74.8 | **76.8** | *77.7* | **0.818** | *0.844* |

Accuracy: Balanced Accuracy
Emphasized in italics are the values where LLM outperforms RF in Accuracy and AUCROC

Our model demonstrates several improvements over random forest for predicting neurodevelopmental delays with the model using Bayley scores at 22 months. Specifically, it achieves a 4.2% increase in AUCROC in MDI prediction and a 3.2% increase in PDI prediction, compared to random forest. Our model outperforms the random forest-based model in sensitivity overall, indicating its superior ability to detect infants at risk, which is critical for timely interventions. Our model strikes a better balance between sensitivity and specificity, highlighting its ability to predict both delayed and non-delayed infants effectively. Moreover, our Model 8, using the scores measured at 19 months of age, outperforms the random forest-based model in 22 months in MDI prediction.

To further assess the significance of our model’s performance compared to the random forest model, we conducted 7-fold cross-validation and paired t-test on Model 9, which uses maternal and perinatal records and longitudinal data from 22 months. As shown in Table 5, our model outperforms the random forest model in terms of accuracy. Moreover, the improvement in sensitivity for mental development delay is also statistically significant.

**Table 5:** Performance comparison for Model 9 and random forest model.

| Model(22 mo) | Mental Development (MDI) |  |  |  | Psychomotor Development (PDI) |  |  |  |
| --- | --- | --- | --- | --- | --- | --- | --- | --- |
|  | Sensitivity | Specificity | Balanced Accuracy | AUCROC | Sensitivity | Specificity | Balanced Accuracy | AUCROC |
| RF | 72.07 ± 10.26** | 86.44 ± 3.70 | 79.24 ± 3.94 | 0.862 ± 0.035* | 78.24 ± 5.70 | 73.84 ± 5.07 | 76.04 ± 2.89** | 0.825 ± 0.031 |
| LLM | **77.50 ± 8.86** | 84.94 ± 5.45 | **81.21 ± 3.88** | **0.879 ± 0.040** | 80.97 ± 8.04 | 75.70 ± 5.17 | **77.69 ± 2.87** | **0.833 ± 0.025** |
| *p-value* | 0.006 | 0.171 | 0.056 | 0.012 | 0.164 | 0.498 | 0.003 | 0.166 |

\*: $p < 0.05$ \*\*: $p < 0.01$

To evaluate the effectiveness of each component of our model (see “Methods” section for detailed description of each component), we conducted an ablation study just for Model 1 and Model 9. Table 6 demonstrates that our full model performs best among all models in predicting target features overall. Even paraphrases generated using a simple prompt achieve better performance compared to key-value pairs. This highlights the LLM’s capability to incorporate domain-specific information from feature names to supplement descriptions of values in tabular data. Further performance gains are achieved by augmenting the dataset through generating five different paraphrases per row. One-shot learning combined with additional instructions also help the LLM capture inter-feature logic, which is proved crucial in achieving a performance level that surpasses the random forest-based model. Lastly, excluding not-observed attributes from binary columns indeed helps the LLMs focus on the more important features for each individual infant and generate better description.

**Table 6:** Ablation table for Model 1 and 9.

| Model | MDI at birth |  | PDI at birth |  | MDI at 22 months |  | PDI at 22 months |  |
| --- | --- | --- | --- | --- | --- | --- | --- | --- |
|  | Accuracy | AUCROC | Accuracy | AUCROC | Accuracy | AUCROC | Accuracy | AUCROC |
| Lin | 53.2 | 0.520 | 63.9 | 0.689 | 77.2 | 0.808 | 71.5 | 0.740 |
| Simp-P | 58.4 | 0.607 | 64.5 | 0.701 | 78.9 | 0.819 | 76.2 | 0.831 |
| Simp-5P | 63.1 | 0.653 | **66.5** | 0.705 | 81.6 | 0.831 | 77.4 | 0.833 |
| 1S-5P-AllFeat | 61.8 | 0.643 | **66.5** | 0.697 | 82.3 | 0.907 | 77.2 | 0.831 |
| **1S-5P (Ours)** | **65.1** | **0.748** | **63.7** | **0.726** | **84.2** | **0.915** | **77.7** | **0.844** |

Lin: linearization key-value pairs as input for BioBERT.
Simp-P: simple prompt + one row one paraphrase.
Simp-5P: simple prompt + one row five paraphrases.
1S-5P-AllFeat: our model but not excluding not-observed attributes in linearization.

### Feature analysis

To understand how our fine-tuned BioBERT model makes its predictions, we employed Shapley Additive exPlanations (SHAP) [27]. SHAP is a powerful technique from game theory that explains a model’s output by quantifying the contribution of each feature to the prediction. However, a standard feature-level SHAP analysis is not applicable for transformer-based models like BioBERT, whose fundamental features are tokens. To overcome this limitation, we used TransSHAP [28], a method specifically designed for applying SHAP to transformer models.

Analyzing tokens in isolation can destroy the meaning of critical multi-word phrases. For example, the clinical concept “intrauterine growth restriction” is only meaningful when the words are bundled together. A standard token-level analysis would incorrectly treat “intrauterine,” “growth,” and “restriction” as independent features, failing to capture their combined importance. To ensure correct interpretation of complete concepts, we identified important multi-word phrases using Pointwise Mutual Information (PMI). PMI evaluates how often tokens co-occur compared to how often they would be expected to appear together by chance. A higher PMI score indicates a stronger semantic bond between tokens. We calculated the PMI scores for bigrams and trigrams across our training data and fused the phrases with high PMI scores into single tokens. For example, the three separate tokens [‘intrauterine’, ‘growth’, ‘restriction’] are fused into a single token: ‘intrauterine_growth_restriction’. By applying TransSHAP to this pre-processed text, our analysis can calculate a more unified SHAP value for the entire concept.

Our training data contains a high frequency and wide variety of numerical values. These different digits often rank prominently in SHAP value analyses, even after fusing key phrases. We made a deliberate decision to filter out all digits before conducting our final analysis. This approach allows us to gain a much clearer and more direct understanding of how our fine-tuned BioBERT model leverages textual features to make its decisions.

After filtering the synonym phrases, Figure 3 displays the top eight phrases that influence Model 1’s predictions, ranked by their SHAP values. The results are shown for both the MDI and PDI, corresponding to delay and no delay outcomes. To enhance clarity and interpretability, terms indicating multiple births like “triplets” and “quadruplets” are consolidated into a single “multi born” feature. Moreover, vague phrases like “severe abnormality” are clarified by appending more descriptive contextual terms in parentheses. We have also done the same feature analysis for our Model 9. The main difference is that the description for the Bayley score also ranks high in the SHAP summary plot for Model 9.

**Figure 3.**
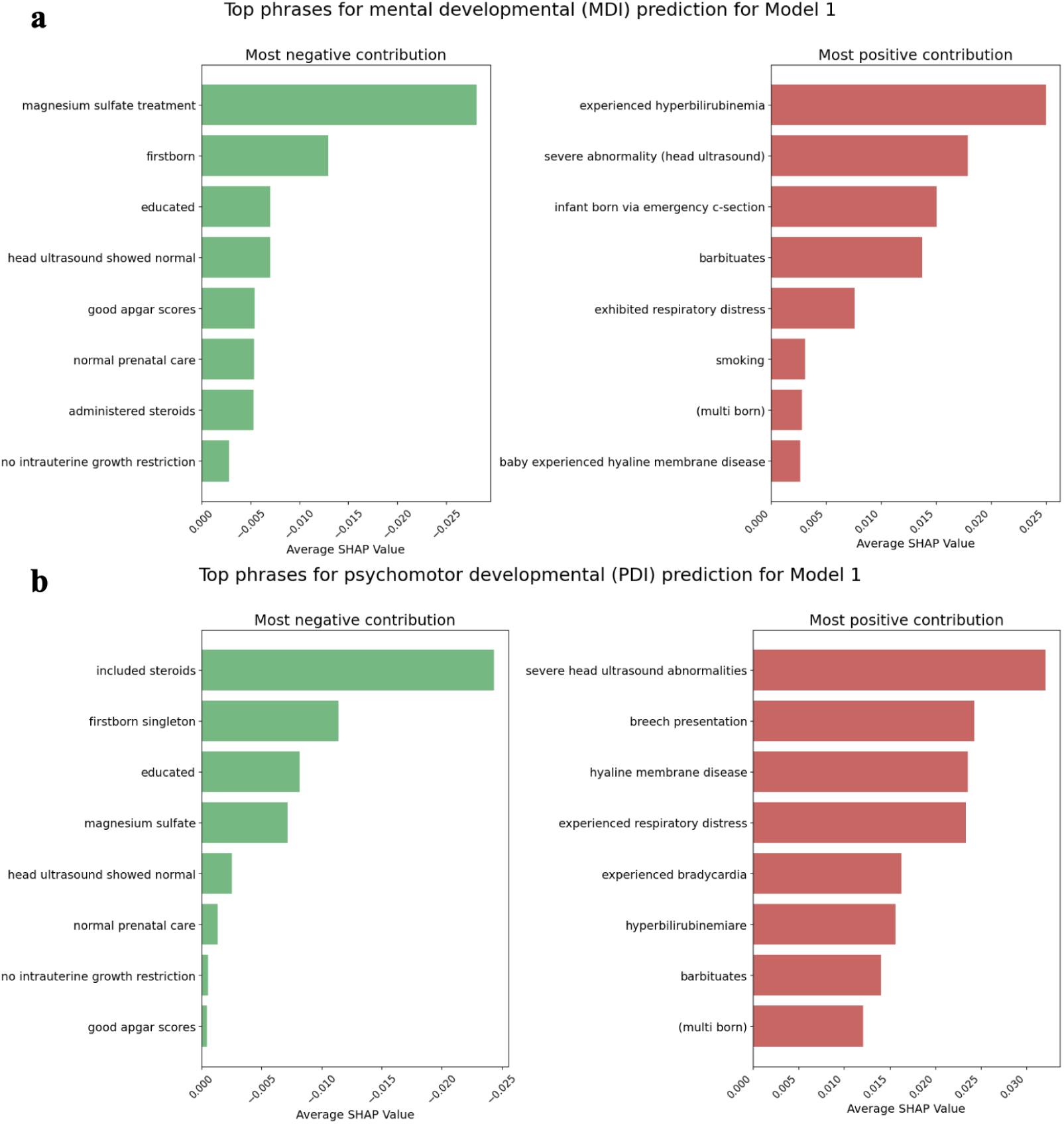
SHAP summary plot for Model 1. A positive SHAP value (x-axis) indicates a higher likelihood of a delay.

## Discussion

In this study, we successfully used LLMs to predict the mental and psychomotor development of very preterm infants using maternal and perinatal records at birth and longitudinal features up to 22 months. Among the models evaluated, the one incorporating MDI and PDI scores at 22 months achieved the highest balanced accuracy in predicting neurodevelopmental delays at 25 months, surpassing the performance of traditional machine learning models. Our model also achieved higher sensitivity. These results show that our model has the superior ability to detect infants at risk of cognitive/verbal and fine/gross motor delay 3 months earlier, which is very helpful for the timely intervention and optimizing their outcomes. Furthermore, our model predicting cognitive/verbal risk 6 months in advance achieved performance nearly as strong as our 3 months later model. It also outperforms 3 months later random forest-based model. These results effectively double the window for early detection and intervention, while maintaining high performance. Juul et al. [29] also utilized perinatal and maternal data, employing Bayesian Additive Regression Trees (BART) to predict neurodevelopmental delay in extremely preterm infants at 2 years of age. They reported a 78.5% balanced accuracy and an AUCROC of 0.871 using only features measured at birth, compatible to our Model 1. A potential reason for their higher accuracy is their stricter classification of delay (Bayley score below 70), which makes the groups easier to distinguish. Bowe et al. [6] employed a data preprocessing approach very similar to ours, also using random forest for missing data imputation and SMOTE for minority class augmentation. Their model achieved a slightly better balanced accuracy (69%) and AUCROC (0.77) than our Model 1. Nevertheless, their dataset had a lower missing data rate than ours–they retained features available in over 75% of the pre-terms, while we set the threshold to 20%. Our less favorable comparison to earlier studies [6, 29] likely also due to our cohort’s composition. Unlike previous datasets spanning about five years, ours covers an 18-year period. This broader time frame introduces greater heterogeneity due to medical advancements, making classification more challenging.

Furthermore, all these approaches were fully supervised, as they had no missing target variables. In contrast, our dataset includes data with some missing target variables, making our training semi-supervised. Keeping data with missing target variable is crucial to retain the maximum amount of data for model training for higher generalizability, especially for studies with relatively small datasets. Additionally, we incorporate longitudinal data, allowing for an analysis of infant developmental trends not captured in these prior studies.

There are also studies incorporating longitudinal data to the prediction. Chung et al. [30] incorporated longitudinal data in 6 and 12 months with other perinatal and maternal data to predict if the infants’ mental and psychomotor Bayley scores below 85. They achieved the balanced accuracy of 69.8% and 73.5% and the AUCROC of 0.78 and 0.82 in predicting mental and psychomotor delay. The reason for slightly better performance compared to our Model 6 is similar to [6, 29]. Using the same dataset, Gozde et al. employed a random forest model to predict neurodevelopmental delay at 25 months of age, based on data from 6 months prior [17]. They achieved the balanced accuracy of 71.7% in MDI prediction. Our Model 8 in MDI outperformed their study. These studies used machine learning based models instead of LLMs, which overlook the incorporation of domain-specific information derived from feature names and large-scale pre-training. Our model leverages the multi-step reasoning ability of LLMs with the pre-trained domain-specific knowledge to capture deeper inter-feature logic.

Consistent with other machine learning approaches [31], our fine-tuned BioBERT classifier identified many features like magnesium sulfate administration, cranial ultrasound, and maternal education as key predictive features. Our SHAP analysis also confirmed the previous findings from machine learning models [17] that prenatal care can potentially prevent the neurodevelopmental delay of preterm infants, although we recognize this can be a proxy for broader socioeconomic variables, especially where healthcare access is unequal. However, compared to traditional machine learning models, our BioBERT classifier pays more attention to specific neonatal pathologies like hyaline membrane disease, respiratory distress, and hyperbilirubinemia. This suggests that BioBERT, by leveraging its deep understanding of biomedical language, identifies the more direct, physiological drivers of neurodevelopmental delay.

There are two main limitations in this study. First, we excluded the digits during the feature analysis, which may have led to an oversight of valuable information regarding our fine-tuned BioBERT model’s predictive behavior. Second, our dataset spans an 18-year period of infant births. It is probable that considerable evolutions in clinical treatment protocols and medical capabilities over this period influenced the model’s predictions, possibly restricting its generalizability to contemporary medical practices.

Future work will focus on two key areas. First, we will incorporate additional data modalities, including audio from diagnostic conversations. Second, we will enrich our existing dataset by adding more detailed descriptions for the cranial ultrasound records. Our ultimate objective is to develop a unified model capable of synthesizing this multimodal information to further enhance predictive accuracy.

Our study highlights the potential of LLMs as powerful tools for assessing the risk of neurodevelopmental delays in preterm infants. This study underscores the importance of integrating state-of-art deep learning techniques into neonatal care, paving the way for more personalized and proactive interventions for at-risk populations, thus opening the door to earlier, AI-driven interventions in neonatal care.

### Ethics statement

Ethics approval was obtained from the IRBs at the Institute for Basic Research in Developmental Disabilities and Richmond University Medical Center. Written informed consent was obtained from parents/guardians for all infants.

## Data availability

The data that support the findings of this study are available from The Research Foundation for Mental Hygiene, but restrictions apply to the availability of these data, which were used under license for the current study and therefore are not publicly available. Data are however available from the authors on reasonable request and with permission of The Research Foundation for Mental Hygiene.

## Acknowledgements

We thank the families who generously allowed their children to participate, the medical staff of Richmond University Medical Center, and the numerous research assistants at the NYS Institute for Basic Research for their generous support.

This study was funded by NICHD grant P01-HD47281, R01-HD21784, PSC-CUNY Award 65406-00 53, and NYS OPWDD. The funder played no role in study design, data collection, analysis and interpretation of data, or the writing of this manuscript.

